# Food insecurity among adolescent girls who are mothers (10-19 years) in sub-Saharan Africa: A scoping review protocol

**DOI:** 10.1101/2025.05.26.25328158

**Authors:** Suliat Fehintola Akinwande, Carmen H. Logie, Peter A. Newman, Kehinde O. Akinwande, Notisha Massaquoi

## Abstract

**Background:** Adolescent girls in resource-constrained settings are vulnerable to sexual exploitation and gender inequities due to poverty, leading to teen motherhood, which is often characterised by stigmatization. Marginalized adolescent mothers may experience stigma in their family, school, community, health, and public services, resulting in poverty, which exacerbates food insecurity. This scoping review will explore the drivers, coping strategies, and the mental, sexual and reproductive health impacts associated with food insecurity among adolescent mothers (ages 10-19 years) in SSA.

**Methods:** We will apply the scoping review framework outlined by Arksey and O’Malley and further developed by the Joanna Briggs Institute, and the reporting guidelines from the Preferred Reporting Items for Systematic reviews and Meta-Analyses extension for Scoping Reviews (PRISMA-ScR) (S1Table). The overall research question is: What are the drivers, coping strategies, and the mental, sexual and reproductive health impacts associated with food insecurity among adolescent mothers (10-19 years) in SSA? In collaboration with a research librarian, search strategies will be developed using text words and subject headings (eg, Medical Subject Headings (MeSH), Emtree) related to adolescent mothers and food insecurity in SSA. We will include quantitative, qualitative, mixed-methods, and review studies in our analysis. A thematic analysis will be conducted on the findings, with results presented in both narrative and tabular formats.

**Ethics and Dissemination:** Formal ethical approval is not required as we are not collecting primary data. The findings will be published in a peer-reviewed journal and disseminated at international conferences.

## Introduction

Globally, 16 million adolescent girls aged 10-19 give birth annually, with birth rates ranging from 12 to 97 births per 1000 teenage girls in high- and low-income countries, respectively (1). SSA has the highest prevalence of adolescent motherhood (18.8%; 95% CI: 16.7–20.9) compared to global regions (2), and this occurs in the context of significantly high food insecurity, with the region representing 10% of the 3.1 billion global population experiencing chronic hunger (3). Research has revealed that adolescent motherhood is associated with negative health and social outcomes (4,5). The analysis of nationally representative data from 144 countries including SSA countries showed an increased risk of mortality in adolescents (10-19 years) compared to women aged 20–24 years (maternal mortality ratio 260 [95% CI 100–410] *vs* 190 [95%CI 120–260] maternal deaths per 100□000 live births (4). Further, teen mothers are often marginalised and stigmatised as irresponsible, promiscuous, and incompetent girls (6,7). This stigma may be experienced in education, healthcare, community and family support—further exacerbating food insecurity (5).

Food insecurity persists as a major public and social health challenge in SSA (8), with an estimated one in five individuals undernourished (3). The concept of food security is defined as the constant physical and economic access to sufficient, safe and nutritious food to fulfil the dietary requirements for an active and healthy life (9). This indicates that food should be continuously available and accessible throughout the life course. The burden of food insecurity is particularly evident among 41% of children under-five in SSA experiencing stunting, a growth retardation condition caused by chronic nutritional deficiencies (10). Early-life malnutrition occurring within the first 1000 days may result in an increased risk of nutrition-related chronic disease, poor cognitive development, lost productivity and death (11). Food insecurity is also linked to a variety of negative psychosocial and physical health outcomes in adolescents, as this stage of life is particularly sensitive due to active growth and development (12). Relative to older mothers, adolescent girls are more likely to be malnourished and have a low-birth-weight baby, who is more likely to become malnourished and be at increased risk of illness and death than those born to older mothers (13). Hence, the food security of adolescent girls and their children living in resource-limited areas of intersecting poverty, food insecurity, and disease is of urgent concern (14).

Resource Scarcity Theory (RST), developed by Wutich and Brewis (15), examines how individuals experience, respond to, and are affected by limited access to essential resources. RST outlines the key drivers of scarcity across social (e.g., age and social exclusion), economic (e.g., inflation and weak governance), and ecological (e.g., drought and flooding) domains (15). It also highlights the broader consequences of resource scarcity on human well-being, including its impact on mental health (e.g. anxiety), as well as adverse sexual and reproductive health outcomes (e.g. HIV). RST offers a comprehensive framework for understanding food insecurity among adolescent mothers.

A range of factors contribute to food insecurity in SSA, including poverty, climate change, and conflict (16). Poverty is a primary obstacle to accessing food among displaced persons, rural and urban poor communities who are vulnerable to urban-biased distribution, foreign aid dependence, and fluctuations in foreign exchange (16). Secondly, climate change in the form of drought and flash floods has led to crop damage, loss of soil fertility, increased soil toxicity, and disruption of the soil ecosystem in different parts of Africa (17). Conflicts also contribute to food insecurity by limiting food production, distribution, availability, and accessibility (18).

However, adolescent mothers may experience other social and structural barriers due to the stigma of teenage motherhood, which may further impact their food insecurity and, consequently, their children’s malnutrition. Therefore, it is pertinent to explore the drivers of food insecurity among adolescent mothers in SSA.

Strategies to mitigate food insecurity have been developed and assessed mainly in relation to the general population, with limited attention to adolescent mothers facing marginalization and discrimination in SSA (19). These strategies include climate-resilient agricultural development interventions, ranging from community gardens, input supplies, land tenure, market access, and postharvest handling (20). Cash transfers (conditional and unconditional) and income□generation interventions have also been reported to minimize the impact of high food prices due to economic hardship (20). Social support has further been shown to alleviate food insecurity by leveraging networks, trust, and cooperative efforts to enhance food accessibility among women (mean age 28.73±4.41 years) (21). However, there remains a gap in knowledge on coping strategies employed by these marginalised adolescent mothers to mitigate food insecurity.

SSA is home to the world’s fastest-growing adolescent population (ages 10–19), which is projected to reach 435 million by 2050 (22). Driven by poverty, which also fuels food insecurity, adolescent girls often engage in sexual relations, further exacerbating the issue of food insecurity (23) . The vicious cycle continues to predispose teenage girls to sexual relations and early childbearing, resulting in disrupted education, limited job prospects, and increased financial strain, leading back to food insecurity (23). Therefore, the well-being of adolescent mothers and their children in these resource-limited regions is important to break this cycle and achieve the Sustainable Development Goal of ending hunger and improving nutrition, leaving no one behind (24). It is imperative to develop a comprehensive understanding of drivers, coping strategies, and the mental health and sexual and reproductive health impacts associated with food insecurity among adolescent mothers.

Scoping reviews are valuable for identifying relevant literature, mapping available evidence, and identifying knowledge gaps (25). Only one scoping review was identified regarding the experiences, challenges, and efforts to support adolescent mothers and their children living in HIV-affected communities in SSA (26). Toska et al. revealed that adolescent mothers living with HIV experience physical and emotional health challenges and poorer treatment outcomes due to discrimination (26). In addition, they report the issue of marginalization of adolescent mothers due to stigmatisation associated with young motherhood and HIV, which predisposes them and their children to sub-optimal health and well-being (26). Building on this review, the present scoping review addresses knowledge gaps on the drivers, coping strategies, and the mental health and sexual and reproductive health impacts associated with food insecurity among adolescent mothers in SSA.

## Method

We will employ the methodological framework of Arksey and O’Malley, which has been further refined by the Joanna Briggs Institute (27). This framework involves the following stages: i) identifying the research question ii) identifying relevant studies iii) study selection iv) charting the data, and v) collating, summarizing, and reporting the results (27). Each stage is described in detail below in alignment with the objectives of the scoping review, adhering to the Preferred Reporting Items for Systematic Review and Meta-Analysis extension for scoping reviews (PRISMA-ScR) guideline (28).

### I. Identifying the research question

In developing the scoping review question, the Population, Concept and Context (PCC) mnemonic will be used as a guide to ensure clarity and efficiently address the topic of the scoping review. The scoping review protocol is titled “Food insecurity among adolescent mothers (10-19 years) living in sub-Saharan Africa -A scoping review protocol” based on the research question. The protocol title is structured to align with the PCC mnemonic, where P represents the population, specified in this protocol as “adolescent mothers (10-19 years)”. The C represents the concept, which is “food insecurity,” and the C-context is “ sub-Saharan Africa.” The scoping review is guided by the following broad questions:

- What are the drivers of food insecurity among adolescent mothers (10-24 years) in SSA?
- What coping strategies do adolescent mothers (aged 10–19 years) in Sub-Saharan Africa (SSA) employ in response to food insecurity?
- What are the mental, sexual, and reproductive health impacts of food insecurity among adolescent mothers (10-19 years) in SSA?

### II. Identifying relevant studies

In consultation with a University of Toronto librarian, electronic databases were reviewed for their relevance and scope in covering the topic. After careful assessment, five databases were selected for the search: Medline, Sociological Abstracts, Africa-wide, PsycINFO, and Scopus. Concepts guiding database searches are from three broad terms.

These terms include:

▸ food insecurity (including food scarcity, hunger, famine, malnutrition, undernutrition); AND
▸ Adolescent mother (young mothers, young motherhood, adolescent mom, teen moms, teenage mothers)
▸ sub-Saharan Africa (Angola, Cameroon, Central African Republic, Chad, Congo (Republic of the Congo, Democratic Republic of the Congo (DRC), Equatorial Guinea, Gabon, Sao Tome and Principe, Burundi, Comoros, Djibouti, Eritrea, Ethiopia, Kenya, Madagascar, Malawi, Mauritius, Mozambique, Rwanda, Seychelles, Somalia, South Sudan, Tanzania, Uganda, Zambia, Zimbabwe, Botswana, Eswatini (formerly Swaziland), Lesotho, Namibia, South Africa, Benin, Burkina Faso, Cape Verde, Côte d’Ivoire, Gambia, Ghana, Guinea, Guinea-Bissau, Liberia, Mali, Mauritania, Niger, Nigeria, Senegal, Sierra Leone, Togo)

The Databases will be searched using text words and, wherever possible, subject headings and validated searches. Database search fields to be examined for text words will include title, abstract and author-assigned keywords (wherever possible). A pilot search of Medline will be translated for use in all other databases to ensure consistency across search platforms and in results. An example search string is presented in the S2 Fig. In addition to searching the noted databases, we will also search grey literature to identify any non-indexed literature of relevance to the scoping review following a comprehensive search guide developed by Godin et al (29). The grey literature search will focus on professional organisation publications and reports. Reference lists from identified studies and reports will also be reviewed to identify additional studies that may not have been captured through the initial search. All searches will be restricted to English-language publications and studies in SSA region. However, a 20-year limit range of publication date restrictions will be applied to ensure comprehensive coverage of relevant literature. Quantitative, qualitative, mixed methods or reports will be considered for inclusion. The age band for adolescence, according to WHO’s definition, is the period of life between 10 and 19 years of age. Therefore, articles will be excluded if more than 50% of their participants are outside the age range specified for adolescent mothers (<10 or >19 years). Retrieved records will be managed using Zotero and Covidence (30).

### III. Study Selection

#### Title and Abstract Screening

This process will be conducted by three members of the research team (FA, KA & TI) by applying the PRISMA-ScR standards recommended for peer-reviewed publications (31). Following the removal of duplicates, the reviewers will independently assess all identified titles and abstracts for eligibility. Covidence software will be used for screening and to monitor agreement between the two reviewers’ assessments. Differences will be resolved through discussion and consensus; if consensus cannot be reached, articles will be moved to the full-text screening for further review.

#### Full-text screening

The full-text screening will be completed independently by the three authors (FA, KA & TI) using the same criteria outlined above. Covidence software will be used to conduct the screening and monitor agreement between the reviewers ‘assessments (30). Similarly, differences in screening will be resolved through discussion and consensus, or in consultation with a third reviewer if needed. We will further check the reference lists of articles that meet the criteria for the full-text review for additional articles relevant to the review. The result of the search and study inclusion process will be presented in PRISMA-ScR flow diagram in Fig 1 (28).

Fig 1. PRISMA flow chart

The PRISMA flow chart (Preferred Reporting Items for Systematic Reviews and Meta-Analyses) will guide the study selection process. Initially, records will be identified through searches of the selected databases. Duplicates and irrelevant studies will be removed based on titles and abstracts (32). The remaining records will then be screened against predefined eligibility criteria to determine their inclusion in the scoping review (32).

### IV. Data charting

We will extract data on publication characteristics, including author(s), month/year of publication, study methods (qualitative, quantitative, or mixed methods), study sites, sample size, and participant demographics. We will also extract information on the key findings related to drivers of food insecurity (e.g., poverty, conflict), coping mechanisms (e.g., consumption adjustment), and impact on well-being among adolescent mothers.

### V. Collating, summarizing, and reporting the results

Two of the reviewers (SA & TI) will extract data from each article and record the results into a Microsoft Excel spreadsheet. Analysis will be confined to a descriptive level as the aim is to map out existing studies on the topic under review. We will then conduct thematic analyses of the findings from the identified studies according to RST (i.e. drivers, coping mechanisms, and health impacts (mental and sexual and reproductive health) linked to food insecurity, in addition to disaggregating findings by country (33). Where relevant, we will also identify intersectional identities (e.g., HIV status, young and single motherhood), how these are addressed in the studies, and their relationship to food insecurity. Data will be presented in narrative form. Additionally, we will chart study limitations or challenges as noted by the study authors, along with the main findings and stated implications.

#### Patient and public involvement

There will be no patient or public involvement in this research.

#### Ethics and dissemination

This review does not require ethics approval because only readily available resources will be synthesised and presented. The result will be published in peer-reviewed journals and presented at international conferences.

## Discussion

This review will provide a thematic synthesis of the existing literature on the drivers, coping strategies, and the mental and sexual and reproductive health impacts associated with food insecurity among adolescent mothers (aged 10-19 years) in SSA. By mapping the current literature, this scoping review will aim to identify knowledge gaps and inform future research and policy priorities in SSA. Additionally, this study may generate new insights into the underexplored intersections between stressors, coping mechanisms, and impacts on well-being associated with food insecurity among adolescent mothers.

### Strengths and limitations

Researchers may differ in their definition of the adolescent age range and may choose not to engage children under 18 in studies because of stringent ethical approval processes, hence limiting the participation of adolescent mothers. Also, the selected databases may not index some relevant peer-reviewed articles; but to address this, we will manually search the reference lists of included articles for potentially relevant sources.

Despite the potential limitations, to our knowledge, this is the first scoping review to examine the experience of adolescent mothers in SSA in relation to the drivers, coping strategies, and the mental as well as sexual and reproductive health impacts associated with food insecurity among adolescent mothers (aged 10-19 years) in SSA. To ensure rigor, the screening and consensus process will be supported by ongoing team meetings to resolve any conflicts in inclusion or exclusion decisions.

### Current study status

A search strategy was developed on the Medline database in partnership with a librarian (YL) from the University of Toronto, from which 51 literature sources were generated. The search strategy will be adapted for each of the other four databases. The scoping review process and consultations began in February 2025. The results are anticipated to be submitted for publication in November 2025.

## Data Availability

No datasets were generated or analysed during the current study. All relevant data from this study will be made available upon study completion

## Acknowledgement

We would like to thank Yonhee Lee, University of Toronto Research Librarian, for assistance in designing the preliminary literature search strategy

