## Supplementary material for "Food insecurity among adolescent girls who are mothers (10-19 years) in sub-Saharan Africa: A scoping review protocol": S2 fig

### S2 Fig. Draft of Medline search string

Ovid® | My Account | Questions? | Support & Training | Help | Feedback | Log Off

Search | Journals | Books | Multimedia | My Workspace | EBP Tools | What's New

Search History (70) — View Saved

| # ▲ | Searches | Results | Type | Actions | | Annotations |
|---|---|---|---|---|---|---|
| 1 | exp Food Insecurity/ | 2453 | Advanced | Display Results | More | |
| 2 | food insecur*.mp. | 10945 | Advanced | Display Results | More | |
| 3 | food secur*.mp. | 18666 | Advanced | Display Results | More | |
| 4 | hunger.mp. | 15682 | Advanced | Display Results | More | |
| 5 | malnutrition.mp. | 68066 | Advanced | Display Results | More | |
| 6 | undernutrition.mp. | 10504 | Advanced | Display Results | More | |
| 7 | famine.mp. | 2958 | Advanced | Display Results | More | |
| 8 | food scarcity.mp. | 463 | Advanced | Display Results | More | |
| 9 | 1 or 2 or 3 or 4 or 5 or 6 or 7 or 8 | 113006 | Advanced | Display Results | More | |
| 10 | exp Adolescent Mothers/ | 100 | Advanced | Display Results | More | |
| 11 | Adolescent mother*.mp. | 2009 | Advanced | Display Results | More | |
| 12 | young mother*.mp. | 1672 | Advanced | Display Results | More | |
| 13 | adolescent mom.mp. | 0 | Advanced | Save | More | |
| 14 | teen* mom.mp. | 5 | Advanced | Display Results | More | |
| 15 | teen* mother*.mp. | 1482 | Advanced | Display Results | More | |
| 16 | adolescent pregnancy.mp. | 3168 | Advanced | Display Results | More | |
| 17 | early motherhood.mp. | 277 | Advanced | Display Results | More | |
| 18 | 10 or 11 or 12 or 13 or 14 or 15 or 16 or 17 | 7316 | Advanced | Display Results | More | |
| 19 | exp "Africa South of the Sahara"/ | 281575 | Advanced | Display Results | More | |
| 20 | Angola.mp. | 2175 | Advanced | Display Results | More | |
| 21 | Cameroon.mp. | 10818 | Advanced | Display Results | More | |
| 22 | Central African Republic.mp. | 1466 | Advanced | Display Results | More | |
| 23 | Chad.mp. | 1855 | Advanced | Display Results | More | |
| 24 | Congo.mp. | 21491 | Advanced | Display Results | More | |
| 25 | Republic of the Congo.mp. | 8284 | Advanced | Display Results | More | |
| 26 | Democratic Republic of the Congo.mp. | 7678 | Advanced | Display Results | More | |
| 27 | Equatorial Guinea.mp. | 632 | Advanced | Display Results | More | |
| 28 | Gabon.mp. | 2649 | Advanced | Display Results | More | |
| 29 | (Sao Tome and Principe).mp. | 253 | Advanced | Display Results | More | |
| 30 | Burundi.mp. | 1352 | Advanced | Display Results | More | |
| 31 | Comoros.mp. | 711 | Advanced | Display Results | More | |
| 32 | Djibouti.mp. | 562 | Advanced | Display Results | More | |
| 33 | Eritrea.mp. | 914 | Advanced | Display Results | More | |
| 34 | Ethiopia.mp. | 37458 | Advanced | Display Results | More | |
| 35 | Kenya.mp. | 30025 | Advanced | Display Results | More | |
| 36 | Madagascar.mp. | 6898 | Advanced | Display Results | More | |
| 37 | Malawi.mp. | 10960 | Advanced | Display Results | More | |
| 38 | Mauritius.mp. | 1396 | Advanced | Display Results | More | |
| 39 | Mozambique.mp. | 5582 | Advanced | Display Results | More | |
| 40 | Rwanda.mp. | 5479 | Advanced | Display Results | More | |

| | | | | | |
|---|---|---|---|---|---|
| 41 | Seychelles.mp. | 1009 | Advanced | Display Results | More |
| 42 | Somalia.mp. | 3282 | Advanced | Display Results | More |
| 43 | South Sudan.mp. | 986 | Advanced | Display Results | More |
| 44 | Tanzania.mp. | 21409 | Advanced | Display Results | More |
| 45 | Uganda.mp. | 24694 | Advanced | Display Results | More |
| 46 | Zambia.mp. | 8701 | Advanced | Display Results | More |
| 47 | Zimbabwe.mp. | 9675 | Advanced | Display Results | More |
| 48 | Eswatini.mp. | 1031 | Advanced | Display Results | More |
| 49 | Lesotho.mp. | 1182 | Advanced | Display Results | More |
| 50 | Namibia.mp. | 2617 | Advanced | Display Results | More |
| 51 | South Arica.mp. | 4 | Advanced | Display Results | More |
| 52 | Benin.mp. | 5144 | Advanced | Display Results | More |
| 53 | Burkina Faso.mp. | 6380 | Advanced | Display Results | More |
| 54 | Cape Verde.mp. | 686 | Advanced | Display Results | More |
| 55 | Cote d Ivoire.mp. | 15 | Advanced | Display Results | More |
| 56 | Gambia.mp. | 3940 | Advanced | Display Results | More |
| 57 | Ghana.mp. | 19503 | Advanced | Display Results | More |
| 58 | Guinea.mp. | 167760 | Advanced | Display Results | More |
| 59 | Guinea-Bissau.mp. | 1485 | Advanced | Display Results | More |
| 60 | Liberia.mp. | 2467 | Advanced | Display Results | More |
| 61 | Mali.mp. | 5552 | Advanced | Display Results | More |
| 62 | Mauritania.mp. | 952 | Advanced | Display Results | More |
| 63 | Niger.mp. | 17436 | Advanced | Display Results | More |
| 64 | Nigeria.mp. | 50591 | Advanced | Display Results | More |
| 65 | Senegal.mp. | 9261 | Advanced | Display Results | More |
| 66 | Sierra Leone.mp. | 3576 | Advanced | Display Results | More |
| 67 | Togo.mp. | 2236 | Advanced | Display Results | More |
| 68 | 19 or 20 or 21 or 22 or 23 or 24 or 25 or 26 or 27 or 28 or 29 or 30 or 31 or 32 or 33 or 34 or 35 or 36 or 37 or 38 or 39 or 40 or 41 or 42 or 43 or 44 or 45 or 46 or 47 or 48 or 49 or 50 or 51 or 52 or 53 or 54 or 55 or 56 or 57 or 58 or 59 or 60 or 61 or 62 or 63 or 64 or 65 or 66 or 67 | 543252 | Advanced | Display Results | More |
| 69 | 9 and 18 and 68 | 59 | Advanced | Display Results | More |
| 70 | limit 69 to (yr="2005 -Current" and english and last 20 years) | 51 | Advanced | Display Results | More |
